# Assessing GPT-4’s Diagnostic Accuracy with Darker Skin Tones: Underperformance and Implications

**DOI:** 10.1101/2024.04.17.24305928

**Authors:** Edgar Akuffo-Addo, Luna Samman, Leena Munawar, Maya Akbik, Nelly Kokikian, Raquel Wescott, Jashin J. Wu

**Author notes:** **Corresponding Author:** Edgar Akuffo-Addo, 1 King’s College Circle, Toronto, ON M5S 1A8. Edgar Akuffo-Addo and Luna Samman have contributed equally to this study and should be considered as co-first authors. **Competing Interests:** Dr. Wu declares being an investigator, consultant, or speaker for AbbVie, Almirall, Amgen, Arcutis, Aristea Therapeutics, Bausch Health, Boehringer Ingelheim, Bristol Myers Squibb, Dermavant, DermTech, Dr Reddy’s Laboratories, Eli Lilly, EPI Health, Galderma, Janssen, LEO Pharma, Mindera, Novartis, Pfizer, Regeneron, Samsung Bioepis, Sanofi Genzyme, Solius, Sun Pharmaceutical Industries, UCB, and Zerigo Health. **Data Sharing Statement:** The data that support the findings of this study are available from the corresponding author upon reasonable request.

## Abstract

**Introduction:** Conversational artificial intelligence (AI) language models like ChatGPT have emerged as promising tools for patients seeking medical information and guidance. However, their use raises ethical concerns due to the potential for inaccurate medical advice that could harm patients. Previous studies in dermatological machine-learning have highlighted that the underrepresentation of diverse skin types in research could lead to bias and reduced performance in evaluating skin lesions in darker skin tones. This study aims to assess the accuracy of GPT-4 in generating appropriate differential diagnoses and arriving at the correct diagnoses for common skin lesions. Additionally, we investigate any differences in its diagnostic accuracy between darker and lighter skin tones.

**Method:** Fifty images were randomly selected from the Fitzpatrick 17k dataset, a publicly available online collection of clinical images labelled with the appropriate diagnoses and skin types based on the Fitzpatrick scoring system. Half of the images selected represented darker skin tones, Fitzpatrick IV-VI, and the other half represented lighter skin tones, Fitzpatrick I-II. For each selected dermatological condition, GPT-4 was presented with pairs of images - one from a lighter skin tone and another from a darker skin tone. GPT-4 was then asked to provide its top three differential diagnoses and a final diagnosis for each pair. The responses generated by GPT-4 were transcribed and compared against the labels provided in the dataset to evaluate accuracy. Subsequently, a univariate linear regression analysis was conducted to investigate the relationship between Fitzpatrick skin type and diagnostic accuracy of GPT-4.

**Results:** Out of the 50 selected images, the distribution of Fitzpatrick skin types was as follows: 40% were Fitzpatrick type I, 10% were type II, 4% were type IV, 26% were type V, and 20% were type VI. Overall, GPT-4 correctly diagnosed the condition in 28% of the images (n=14/50), while the correct diagnosis was included in its list of top differentials for 48% of the images (n=24/50). GPT-4 exhibited better performance in providing the correct diagnosis for lighter skin tones (44%, n=11/25) compared to darker skin tones (12%, n=3/25), and this was statistically significant (p-value < 0.05). Furthermore, with each unit increase in the Fitzpatrick scale, GPT-4s performance decreased by 11.4% in accurately providing a differential diagnosis and by 7.1% in accurately providing the correct diagnosis.

**Conclusion:** GPT-4 exhibited significantly lower overall accuracy compared to previous studies reporting accuracies as high as 90%. This discrepancy highlights GPT-4s potential limitations in providing accurate information without sufficient clinical context. While GPT-4 could serve as a valuable learning tool for medical students and dermatology residents, it may not be suitable for patients seeking clinical input to self-diagnose lesions at home. It is important to note that this study is limited by its relatively small sample size, which could impact the generalizability of the findings. If GPT-4 is to be considered for use by patients in a clinical setting, it is important to ensure that it demonstrates high accuracy and remains unbiased across all patient demographics and skin types.

## Research letter

Conversational artificial intelligence (AI) language models like ChatGPT have emerged as promising tools for patients seeking medical information and guidance.^1^ However, their use raises ethical concerns due to the potential for inaccurate medical advice that could harm patients.^2^ Previous studies in dermatological machine-learning have highlighted that the underrepresentation of diverse skin types in research could lead to bias and reduced performance in evaluating skin lesions in darker skin tones.^3^ This study aims to assess the accuracy of GPT-4 in generating appropriate differential diagnoses and arriving at the correct diagnoses for common skin lesions. Additionally, we investigate any differences in its diagnostic accuracy between darker and lighter skin tones.

Fifty images were randomly selected from the *Fitzpatrick 17k* dataset, a publicly available online collection of clinical images labelled with the appropriate diagnoses and skin types based on the Fitzpatrick scoring system.^4^ Half of the images selected represented darker skin tones, Fitzpatrick IV-VI, and the other half represented lighter skin tones, Fitzpatrick I-II. For each selected dermatological condition, GPT-4 was presented with pairs of images - one from a lighter skin tone and another from a darker skin tone. GPT-4 was then asked to provide its top three differential diagnoses and a final diagnosis for each pair. The responses generated by GPT-4 were transcribed and compared against the labels provided in the dataset to evaluate accuracy. Subsequently, a univariate linear regression analysis was conducted to investigate the relationship between Fitzpatrick skin type and diagnostic accuracy of GPT-4.

Out of the 50 selected images, the distribution of Fitzpatrick skin types was as follows: 40% were Fitzpatrick type I, 10% were type II, 4% were type IV, 26% were type V, and 20% were type VI. Overall, GPT-4 correctly diagnosed the condition in 28% of the images (n=14/50), while the correct diagnosis was included in its list of top differentials for 48% of the images (n=24/50). GPT-4 exhibited better performance in providing the correct diagnosis for lighter skin tones (44%, n=11/25) compared to darker skin tones (12%, n=3/25), and this was statistically significant (p-value < 0.05). Furthermore, with each unit increase in the Fitzpatrick scale, GPT-4’s performance decreased by 11.4% in accurately providing a differential diagnosis and by 7.1% in accurately providing the correct diagnosis.

GPT-4’s exhibited significantly lower overall accuracy compared to previous studies reporting accuracies as high as 90%.^5^ This discrepancy highlights GPT-4’s potential limitations in providing accurate information without sufficient clinical context. While GPT-4 could serve as a valuable learning tool for medical students and dermatology residents, it may not be suitable for patients seeking clinical input to self-diagnose lesions at home. It is important to note that this study is limited by its relatively small sample size, which could impact the generalizability of the findings. If GPT-4 is to be considered for use by patients in a clinical setting, it is important to ensure that it demonstrates high accuracy and remains unbiased across all patient demographics and skin types.

## Data Availability

All data produced in the present study are available upon reasonable request to the authors

## Acknowledgements

None

